# Knowledge, attitude, and practice of ward attendant and housekeeping staffs towards dead body care of COVID-19 patient at tertiary care hospital : A cross sectional study

**DOI:** 10.1101/2022.08.23.22279058

**Authors:** Giriraj Saini, Prasan K Panda, Maneesh Sharma, Mahendra Singh, Ravi Meshram

## Abstract

**Background:** Healthcare workers worked untiringly during entire pandemic period and taken efforts to protect individuals, families and communities in adverse situations with stretched resources. Among health care workers role of ward attendant and housekeeping staffs have been very significant particularly in infection control practices and dead body management. Present study’s aim is to gain an understanding of the knowledge, attitudes, and practices (KAP) of ward attendant and housekeeping staffs towards dead body management.

**Methods:** Hospital-based cross-sectional study design was conducted among ward attendant and housekeeping staffs working in COVID units. A total of 62 participants were selected using simple random sampling technique. Self-administered questionnaire was used to collect data. Binary logistic regression model was used to see association between outcome and independent variables.

**Result:** Present study found mean knowledge, attitude and practice score of participants were 6.1, 49.9 and 12.28 indicates good knowledge, positive attitude and inappropriate practice towards dead body care. Study result also shows that odds of good knowledge were not significantly associated with demographic variables. However, the participants who didn’t receive any training on dead body care were found to have positive attitude towards dead body care(AOR=3.90,95%CI=1.092-13.92), whereas gender (AOR=1.85,95%CI=.430-7.96), working experience in COVID units (AOR=99.5,95%CI=.913-98.8) and educational qualification (AOR=30.33,95%CI=1.5-577) were significantly associated with practice of dead body care of COVID-19 patients.

**Conclusion:** The study found that majority of participants were having good knowledge, positive attitude and inappropriate practice towards dead body care of COVID-19 patients. Hospital administration should conduct regular training of dead body care of COVID-19 patients for all the housekeeping staffs and ward attendant to minimise the risk of exposure to infections and better management of dead bodies.

## Introduction

Novel Corona Virus Disease (COVID-19) rapidly has been transmitted around the world and has become a pandemic, after the first cases reported in Wuhan, China in the December 2019. After a year of transmission in December 2020, the COVID-19 pandemic has affected 215 countries throughout the world, resulted in more than 48 million cases and more than 1.8 million deaths(1).

The first cases of COVID-19 in India were reported on 30 January 2020 in three towns of Kerala, among three Indian medical students who had returned from Wuhan, the epicenter of the pandemic(2). Later India was reported to be in the first position among the South-East Asian countries and fourth position across the globe with over 2 lakhs of confirmed cases and 8498 deaths by mid of 2020 (3) and according to ministry of health family welfare report (MoHFW) total deaths due to COVID-19 were 5.22 lakhs in India(4). Healthcare workers worked tirelessly during entire pandemic period and tried to protect individuals, families and communities in adverse situations with stretched resources. Among health care workers, ward attendant and housekeeping staffs have played a very significant role during COVID-19 pandemic particularly in infection control practices and dead body management. Although COVID death totals remain undetermined in India however, COVID constituted 29% of deaths from June 2020 to July 2021, corresponding to 3.2 Million deaths, of which 2.7 M occurred in April to July 2021. Therefore, burden of caring dead body was much higher among health care workers during entire period of pandemic. Since the role of ward attendant and housekeeping staffs were significant hence it become paramount to understand the knowledge, attitude and practice of ward attendant and housekeeping staffs regarding dead body of care. Literature suggests that lack of knowledge and misunderstandings among HCW’s leads to spread of disease and poor infection prevention practice.. Currently, there is scarce information regarding knowledge, attitude and practice of ward attendant and housekeeping staffs towards dead body care. Therefore, present study was aimed to determine current status of knowledge, attitude and practice towards dead body care of COVID-19 diagnosed patients among ward attendant and housekeeping staffs.

## Material and methods

A hospital based cross-sectional study was conducted from September to October 2020 at tertiary care hospital, Uttarakhand. To implement social distancing in order to avoid the spread of COVID-19, it was not feasible to collect self reported data-based survey hence the investigators have used an online method of data collection. The sample size calculated by Raosoft assuming a response rate of 70%, 95% confidence interval (CI), Z of 1.96, and margin of error of 5%. A further 10% was added to counteract any errors in completing the questionnaires, resulting in a final sample size of 60. All ward attendant and housekeeping staffs working in COVID units were taken as study population in present study. A questionnaire was designed on Google forms, and a link was shared to WhatsApp groups of WA and HKS. The link was also shared personally to WA and HKS in the contact lists of the investigators.

### Measurement and data collection

The structured questionnaire was prepared after reviewing published literatures based on guidelines, reports, and course material regarding emerging respiratory diseases including COVID-19 (5–8). An initial draft of the questionnaire was designed, and subsequently validated in two phases. Firstly, the study instrument was sent to experts from the filed of medicine, nursing and toxicology and were requested to give their expert opinion with respect to its simplicity, relativity and importance. Secondly, a pilot study was conducted on small sample size i.e 8 WA and HKS. Reliability was calculated using SPSS Version 21, and Cronbach’s alpha was 0.79. The questionnaire contains items on socio-demographic profile, Knowledge, attitude and practice related items. Knowledge section comprised 10 items assessed nature of infectivity, equipment, decontamination solution and duration of cleaning of dead body. Each item was having right response labelled as 1 while wrong response was labelled as 0. Total score ranges from 0–8 and a cut off level of < 6 was set for poor knowledge and >6 (75% and above) for good knowledge. Attitude section comprised of 13 items assessing attitude of WA and HKS toward use of PPE, infection control practices, belonging of deceased COVID patients and Risk of getting infection from dead body of COVID patient. Response of each item was recorded on 5-point Likert scale as follows strongly agree (5-point), agree (4-point), neutral (3-point), disagree (2-point), and strongly disagree (1-point). Total score ranges from 13 to 65, with score of >49 (>75%) indicates positive attitude toward dead body care of COVID patients.. Practice section included 18 items regarding wrapping of dead body, solution used to clean the dead body, biomedical waste disposal, use of PPE, placing of tag over dead body other precautionary measures. Each item was responded as yes (1-point) and no (0-point). Practice items total score ranged as 0–18, and a score of >13 demonstrated good practice and a score of < 13 indicates poor practice toward care of dead body of COVID patient.

### Ethical considerations

Ethical clearance was obtained from institute ethical committee vide letter no AIIMS/IEC/20/558 before conducting the study. The study questionnaire contained a consent section that stated the purpose of the study, nature of the survey, study objectives, voluntary participation, declaration of confidentiality and anonymity.

### Statistical analysis

Descriptive and inferential statistics were applied using SPSS Version 21 (IBM Corp.). Chi-squared, independent t test and Fisher exact test were used to find the association of knowledge, attitude and practice with demographic variables. A univariate logistic regression analysis was applied to identify possible determinants of good knowledge, attitude and practice, with results expressed as odds ratio (OR) and 95% CI. P <0.05 was considered to indicate significance in all tests.

## Result

In total, 62 respondents were included in the final analysis out of which 42% were house keeping staffs and, 58% were ward attendant. The mean age of the participants was 24 years and majority (75%)of them belonged to 21-30 years of age group. About 85% of participants were male, 51% having < 2 years of experience and 47% had work experience in COVID designated area of > 81 days. Majority 31 % of participants were educated up to senior secondary level, while 92% of them had previous experience of handling dead body of COVID patient. 52% of participants did not have any experience of handling dead body of SARS, MERSA, Swine flu, NIPAH and EBOLA patients and Majority 52% of participants did not have any training for handing dead body of COVID patient (Table 2).

**Table 1:**
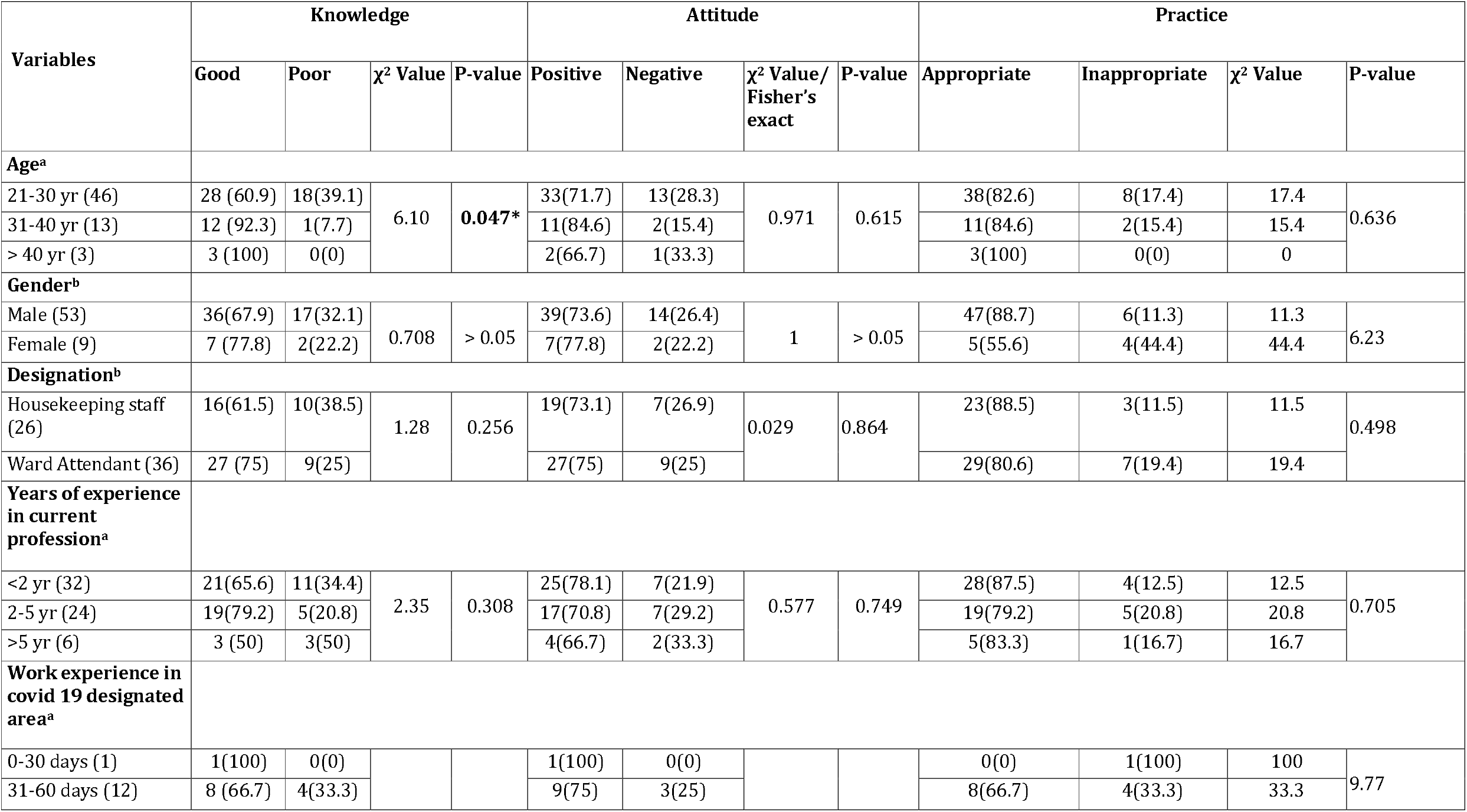
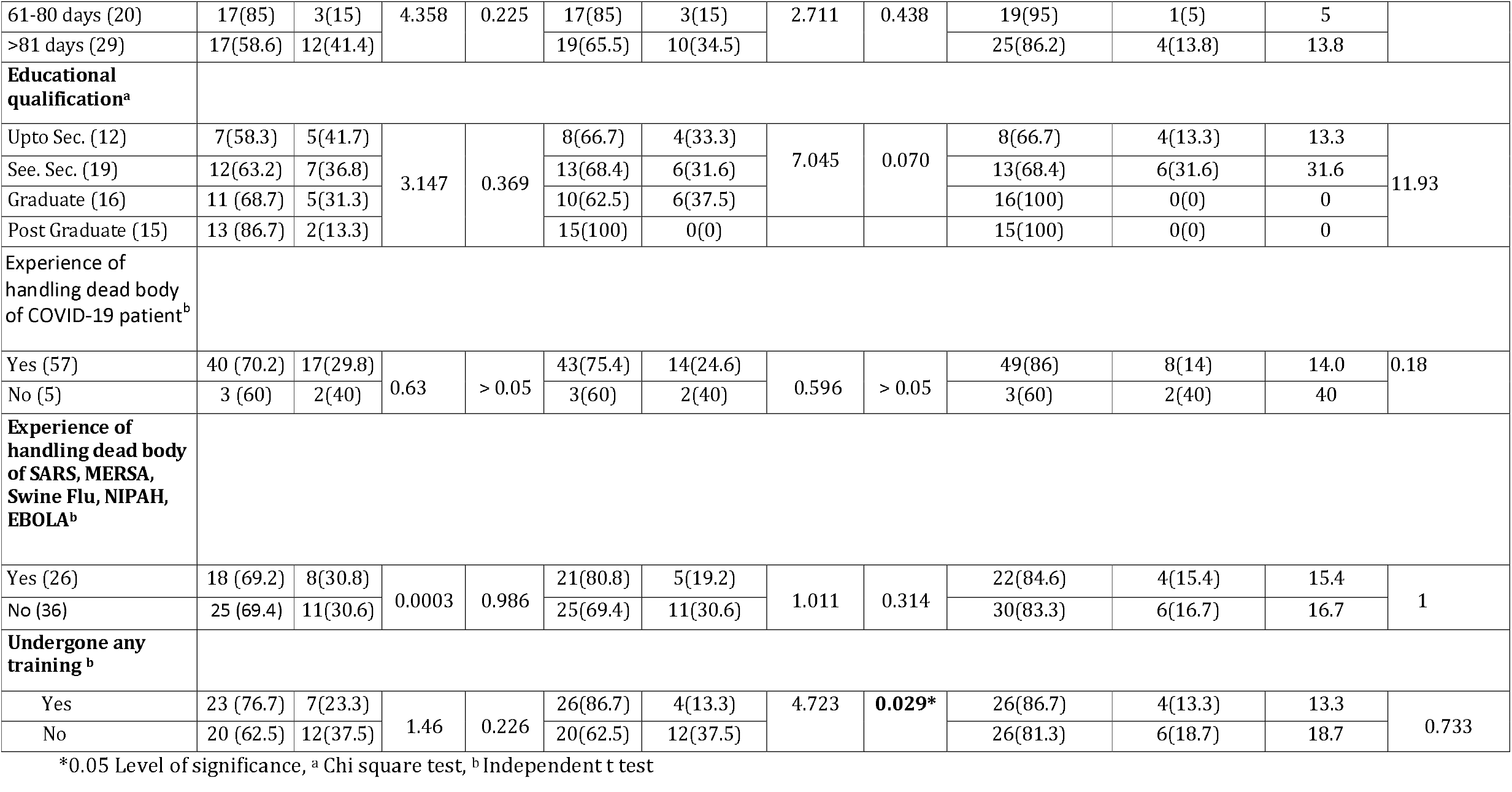
Association of knowledge, attitude and practice with demographic variables (N=62)

**Table 2:**
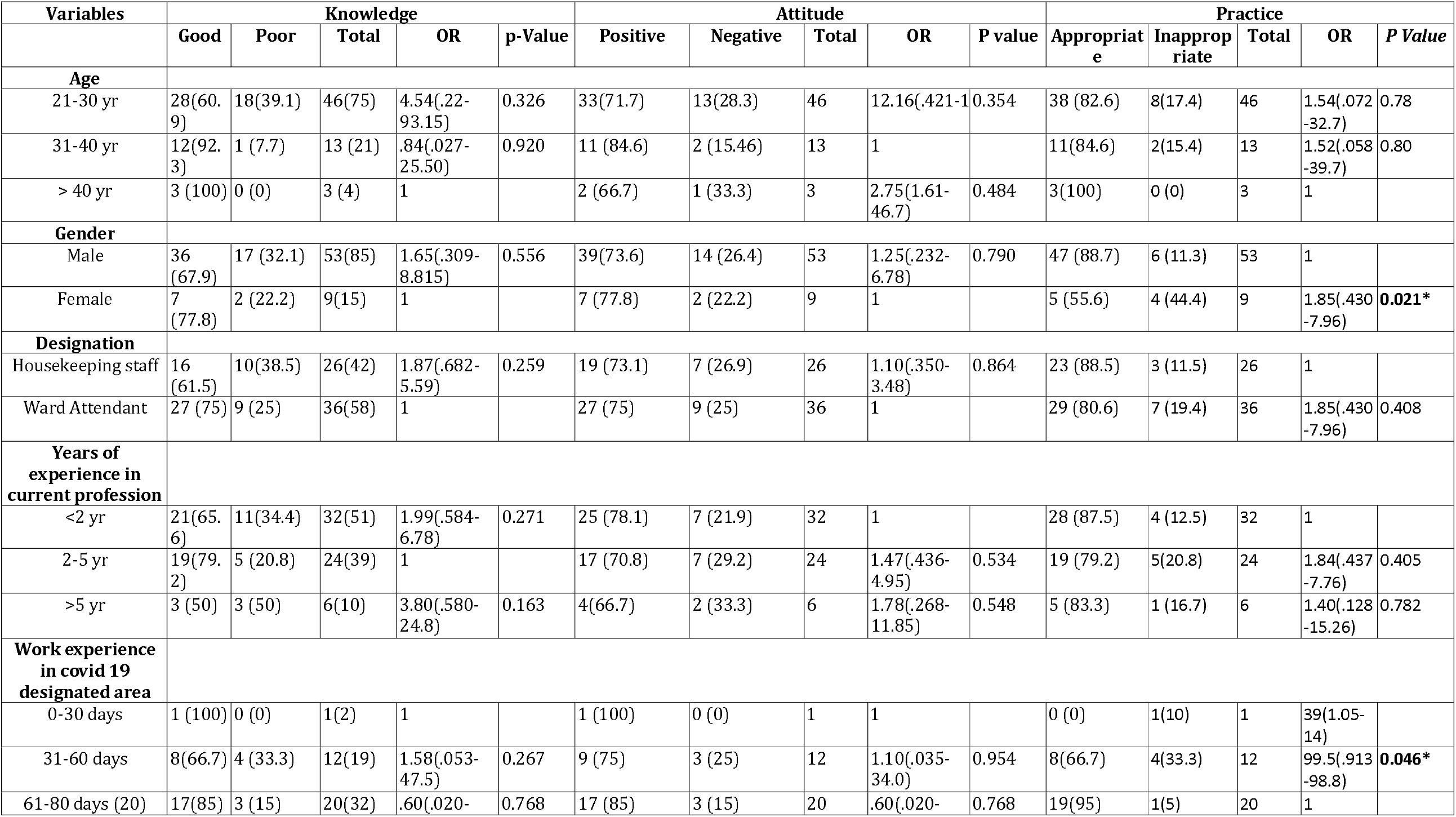
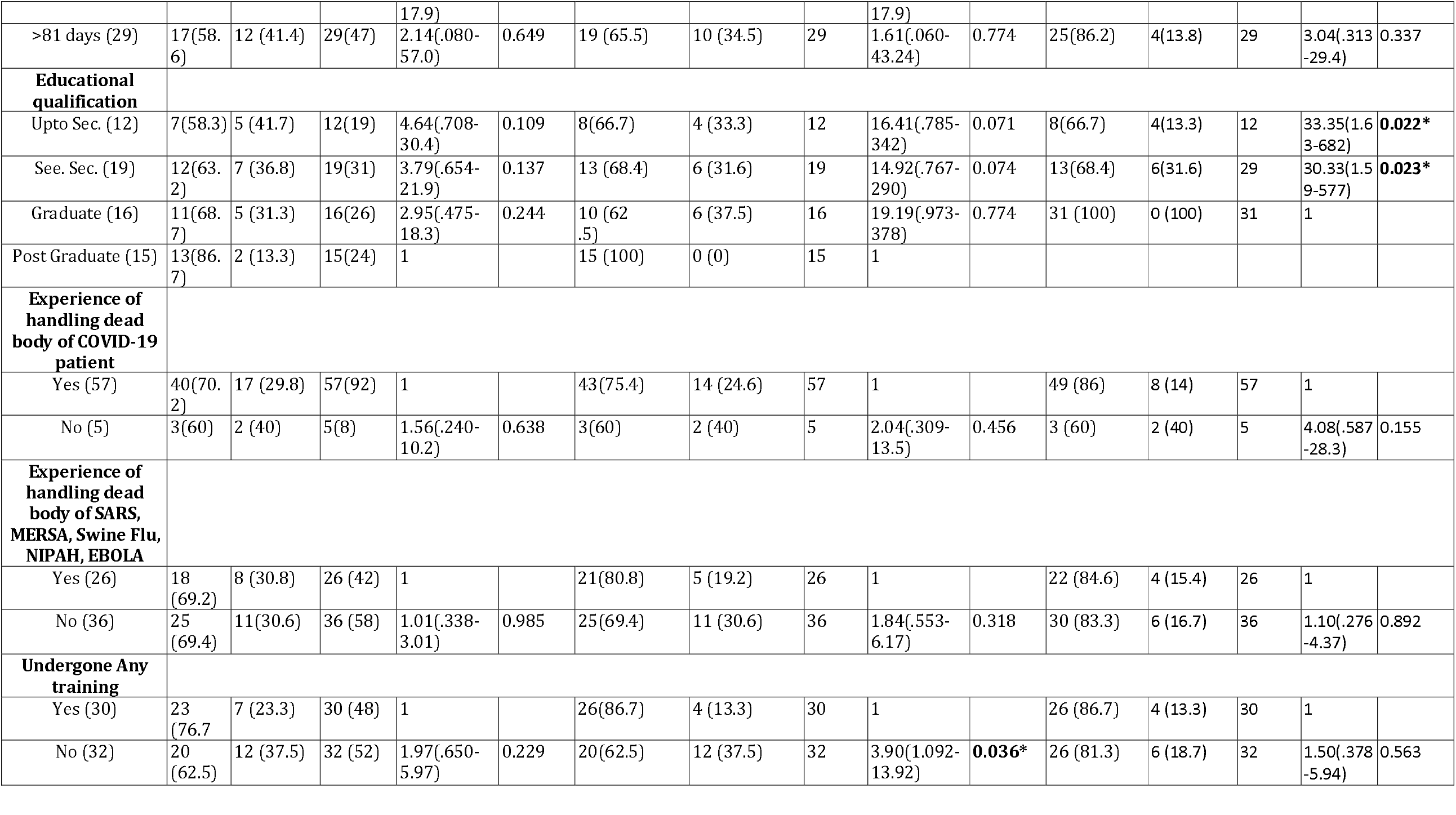
Factors associated with Knowledge, attitude and practice of participants towards dead body care of COVID-19 patient (N=62)

In general, majority of participants showed good knowledge (mean±SD 6.1±1.4), positive attitude (mean±SD 49.9±7.3) and inappropriate practice (mean±SD 12.28±7.1) towards dead body care of COVID-19 patients (Table 3). In association of knowledge, attitude and practice with demographic characteristics age was significantly associated with good knowledge score (p=0.047) and the participants who had undergone training of dead body care were significantly associated with positive attitude (Table 1).

**Table 3:**
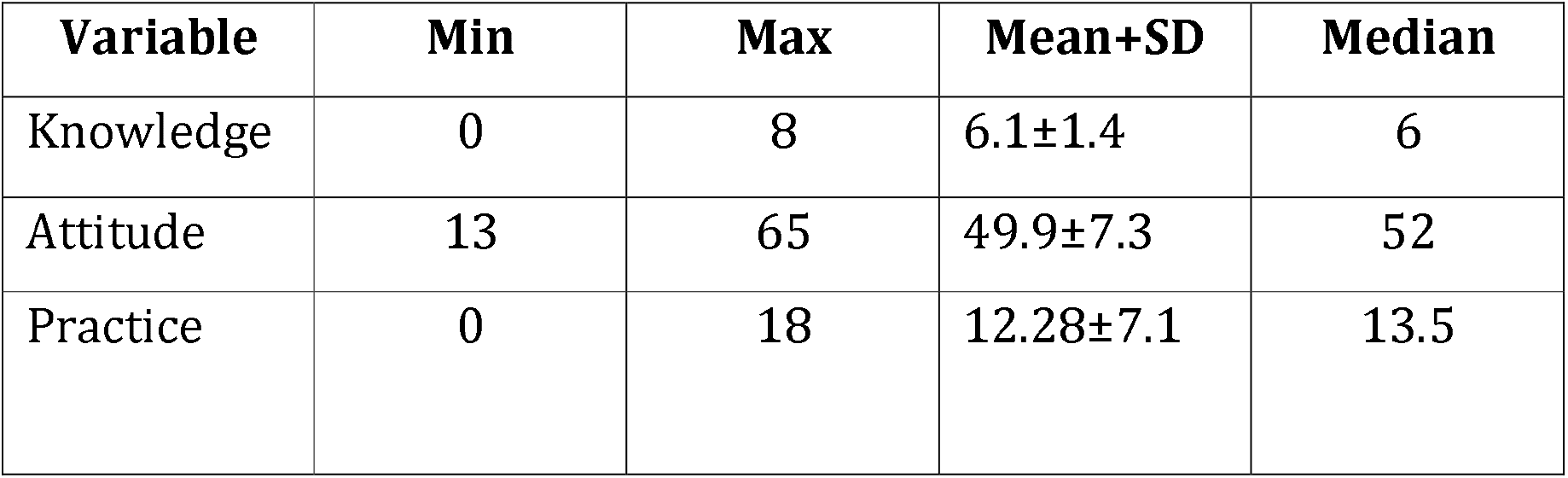
Knowledge, attitude and practice score of participants (N=62)

The output of the logistic regression model odds of good knowledge were not associated with age, gender, designation, years of experience, work experience in COVID units, educational qualification, previous experience of handing COVID 19 patients, experience of handling dead body of SARS, MERSA, swine flu, NIPAH, EBOLA and undergone any training. Further, participants who did not undergone training for dead body care showed significant association and 3.90 times had positive attitude than who did received the training (AOR = 3.90, 95% CI = 1.092-13.92) (Table 2). In regard to practice gender, working experience in COVID units and educational qualification were strongly associated with practice of dead body care of COVID patients. Female participants were having 1.85 more times appropriate practice then male participants (AOR = 1.85, 95% CI = .430-7.96). Participants working between 31-60 days in COVID units had shown more appropriate practice than others (AOR = 99.5, 95% CI = .913-98.8) also participants who were educated upto secondary level (AOR = 33.35, 95% CI = 1.63-682) and senior secondary (AOR = 30.33, 95% CI = 1.5-577) level have shown appropriate practice towards dead body care of COVID patients than those with graduation and above (Table 2).

## Discussion

Present study was conducted to assess the knowledge, attitude and practice of ward attendant and housekeeping staffs towards “Covid-19 dead body care. COVID-19 was an emerging, rapidly changing global health challenge affected everyone including HCW’s. The role of ward attendant and housekeeping staffs were very significant during entire pandemic period particularly in infection control practices and care of dead body of COVID-19 positive patients. Therefore, it was paramount to understand the knowledge, attitude and practice of ward attendant and housekeeping staffs regarding care of dead body of COVID-19 patient. There is paucity of literature on care of dead body of COVID patients and to the best of our knowledge, this is the first study in in India and even in abroad which assess the KAP of ward attendant and housekeeping staffs regarding care of dead body of COVID-19 patient. There were only a single article letter to editor by Ravi KS et al.(9) on dead body management in times of COVID-19 stated that the augmented risk of Covid-19 contamination from a dead body to healthcare workers or relatives who follow standard precautions while handling the body is quite unlikely of COVID patient. In the present study, we were able to demonstrate that about majority of participants were having good knowledge of dead body care of COVID-19 patients and there was no significant association of knowledge with demographic variables except age. In regard to attitude of participants majority were having positive attitude towards dead body care while the participants didn’t undergone any training of dead body care have positive attitude towards dead body care of COVID-19 patients. Present study shows that majority of participants were following inappropriate practice towards dead body care of COVID-19 patients also gender, working experience in COVID units and educational qualification were strongly associated with practice of dead body care of COVID patients. The logistic regression analysis model allowed us to quantify the effect of demographic and knowledge, attitude and practice variables in developing adequate knowledge, positive attitude and appropriate practice towards dead body care of COVID patients. The present study highlighted gender, working experience in COVID designated units and educational qualification as an active predictor for the development of practice outcome towards dead body care of COVID patients as tested in bivariate model. Furthermore, the participants who didn’t undergone any kind of training have shown active predictor for positive attitude towards dead body care of COVID patients.

## Conclusion

In conclusion, we found that majority of ward attendant and house keeping staffs were having good knowledge, favourable attitude and appropriate practice towards care of dead body of COVID-19 patients. We recommend follow up studies including multi-centre and bigger sample size.

## Limitations

Present study was conducted at single tertiary care centre with limited sample size due to COVID constrains.

## Data Availability

All data produced in the present study are available upon reasonable request to the authors

